# Adjusting for time of infection or positive test when estimating the risk of a post-infection outcome in an epidemic

**DOI:** 10.1101/2021.08.13.21262014

**Authors:** Shaun R. Seaman, Tommy Nyberg, Christopher E. Overton, David Pascall, Anne M. Presanis, Daniela De Angelis

## Abstract

When comparing the risk of a post-infection binary outcome, e.g. hospitalisation, for two variants of an infectious pathogen, it is important to adjust for calendar time of infection to avoid the confounding that would occur if the relative incidence of the two variants and the variant-specific risks of the outcome both change over time. Infection time is typically unknown and time of positive test used instead. Likewise, time of positive test may be used instead of infection time when assessing how the risk of the binary outcome changes over calendar time. Here we show that if mean time from infection to positive test is correlated with the outcome, the risk conditional on positive test time depends on whether incidence of infection is increasing or decreasing over calendar time. This complicates interpretation of risk ratios adjusted for positive test time. We also propose a simple sensitivity analysis that indicates how these risk ratios may differ from the risk ratios adjusted for infection time.

## 1 Introduction

Consider the problem of estimating the distribution of time between a first event (e.g. becoming infected with SARS-CoV-2) and a second event (e.g. testing positive for the virus) in the population of individuals who ultimately experience the second event. We shall call this time the ‘inter-event time’ or ‘delay’. Estimating this distribution may be complicated by the first event time not being observed and/or the available data being right-truncated on the second event time, due to only sampling individuals who experience the second event by a particular calendar time.

It has long been known that the inter-event times in the population of individuals who have experienced the second event before a given time tend to be shorter than in the population of all individuals (who eventually experience the second event) [3, 12, 20]. More precisely, the conditional probability that the inter-event time is less than *l* given that the second event has occurred by a given time is greater than the corresponding unconditional probability (unless all second events have occurred by that time).

It has also been noted that the conditional distribution of inter-event time given the actual time of the second event depends on the marginal distribution of the first event time [13, 19, 10]. In particular, if the first event is generated by a Poisson process whose rate is increasing with time, then the conditional distribution of the inter-event time given the second event time is shifted towards zero compared to the unconditional distribution. On the other hand, if the rate is decreasing, the conditional distribution of the inter-event time is shifted away from zero compared to the unconditional distribution. This means that in the context of an infectious disease the time from infection (first event) to positive test (second event) in those who test positive at a given time tends to be shorter than average when the incidence of infection is rising, and longer than average when the incidence is falling.

Now consider a third variable, which is measured at, or after, the time of the second event and is correlated with the inter-event time. Just as the distribution of inter-event time is affected by conditioning on the time of the second event, so might the distribution of this variable. For example, an infected individual’s viral load at time of positive test is a function of time since infection. Rydevik et al. (2016) observed that this relation could be used to estimate an individual’s infection time from that individual’s viral load at the time of testing positive [19]. Hay et al. (2021a) used this same idea to estimate the pattern of incidence of infection in the population from data on the distribution of viral load (measured as the cycle threshold) in a random sample of individuals who tested positive on a given day [10]. If the mean viral load is high, this suggests most of the sampled individuals were infected recently, which is consistent with a rising incidence of infection. Conversely, if the mean viral load is low, this suggests less recent infection, and so a falling incidence. Hay et al. (2021b) investigated using such data to estimate simultaneously the pattern of incidence of infection and the dependence of the viral load on the time since infection [11]. Similar work had previously been done in the field of HIV/AIDS (e.g. [14, 6, 22]).

In the present article we consider the estimation of the distribution of a third variable where this variable is a binary outcome of interest. An association between this binary outcome and the inter-event time could arise due to factors that determine both. We take the first and second events to be infection and positive test, respectively, and the binary outcome to be hospitalisation within 14 days of the positive test, although what follows would apply to any other binary outcome, e.g. death within 28 days of a positive test. Individuals with more severe infections may tend to experience symptom onset sooner after infection — and consequently be tested earlier — than average and also be more likely to become hospitalised. In this situation, the hospitalisation risk (i.e. the proportion ultimately hospitalised) in individuals who test positive before a particular calendar time would be higher than the risk in all individuals who eventually test positive. More importantly for this article, the hospitalisation risk in individuals who test positive *at* a particular calendar time will differ from the risk in all individuals who eventually test positive (unless the incidence of infection is constant over time). If the incidence of infection is rising, the former risk will be higher than the latter; if incidence is falling, it will be lower.

This dependence of the hospitalisation risk on the trajectory of incidence of infection is particularly relevant for any investigation of how the risk is changing over time. Ideally, such an investigation might involve comparing the risks for individuals with different infection times. If, as is likely, infection times are unknown, it would be natural to instead compare the risks for individuals with different positive test times. The difficulty with interpreting this latter comparison is that, as noted above, even if the risk does not vary by the infection time, it will depend on the time of positive test.

Another situation where one might condition on time of positive test is when comparing the risks associated with two variants of a given pathogen, in this case SARS-CoV-2. Here, controlling for (i.e. conditioning on) time of infection would be important, because the ‘exposure’ (i.e. a binary variable for the variant) and the outcome (hospitalisation) may both depend on calendar time. The exposure would depend on time if the ratio of the incidence rates of infection with the two variants varied over time. That would the case if, for example, one variant emerged earlier but the other variant later became dominant. The hospitalisation outcome would depend on time if measures designed to reduce the need for hospitalisation and/or policies on hospital admission changed over time. Failure to control for infection time when comparing the risks of hospitalisation for the two variants would mean comparing the risk in individuals infected with one variant, whose infection times may have been predominantly when pre-hospital treatments were less effective and/or hospital admission more encouraged, with the risk in individuals infected with the other variant, whose infection times were mostly when pre-hospital treatments were better or hospital admission more restricted. If infection times are unknown, it would be natural to control instead for the time of positive test. The difficulty with this approach is that, even if the hospitalisation risk is the same for both variants and does not depend on time of infection, once we condition on time of positive test a variant that has increasing incidence of infection will appear to have a higher risk than a variant that has a decreasing incidence.

Numerous studies have compared the risks of hospitalisation, intensive care unit admission and/or death in individuals infected with two variants of SARS-Cov-2 (either Alpha versus wildtype or Delta versus Alpha), adjusting for time of positive test [2, 4, 5, 8, 9, 15, 16, 21, 23]. In all these studies, the incidence of one variant has been rising while the other has been falling or has been rising at a slower rate. In this article, we describe in detail why and how the conditional risk of hospitalisa-tion given time of positive test depends on the trajectory of incidence of infection, even when the conditional risk given time of infection does not. We also propose an easily implemented method that provides an indication of how much an estimate of the risk conditional on the positive test time might differ from the estimate one would have obtained if it had been possible to condition on the infection time. This method requires the user to specify a range of plausible values for the difference between the mean time from infection to positive test in those individuals who become hospitalised and the mean time in those who do not become hospitalised.

The structure of the article is as follows. Section 2 defines our notation. Section 3 describes why and how the distribution of the delay conditional on the time of positive test depends on the incidence of infection. Section 4 goes on to explain how this dependence affects the conditional risk of hospitalisation given positive test time. In Section 5 we describe our proposed method, which involves conditioning on an alternative proxy of infection time. The performance of this method is studied in Section 6. Censoring of the hospitalisation outcome is discussed in Section 7. We conclude with a discussion in Section 8.

## 2 Notation

We shall consider the population to be everyone who is at risk of infection from some calendar time zero. Time can be measured discretely or continuously. Suppose for now that all infections result in positive tests. In Section 8 we shall discuss the consequences of violation of this assumption. If an individual has two or more separate episodes of infection, we only consider the first episode.

For each individual in the population, let *I* denote the time of infection, and let *L* be the delay (‘*L*’ for ‘lag’) between infection and positive test. Now, *T* = *I* + *L* is the time of positive test. Let *H* equal 1 if the individual is hospitalised within 14 days of positive test, and 0 otherwise. In Section 3, we shall use *X* to denote a covariate or vector of covariates that are fixed from the time of infection. For example, *X* could be *X* = (*V, U*), where *V* is a binary variable indicating which variant of the virus has infected the individual, and *U* is age and ethnicity. In this situation, we might be interested in the odds ratio of hospitalisation associated with *V* adjusted for *U* and, ideally, time of infection.

## 3 Delay distribution conditional on test time

In this section and Section 4, we shall assume, for simplicity, that *L* is independent of *I*. Using Bayes’ Rule, *f*_*L*_(*l* | *T* = *t*), the conditional probability distribution function of the delay given the positive test occurs at time *t*, can be shown to be related to the unconditional probability distribution function *f*_*L*_(*l*) by

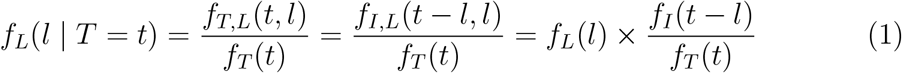

If the incidence of infection is increasing over the period prior to time *t, f*_*I*_(*t* − *l*) will be a decreasing function of *l*. So, for any *l*_1_ < *l*_2_, we have *f*_*I*_(*t* − *l*_1_) > *f*_*I*_(*t*− *l*_2_) and so equation (1) implies

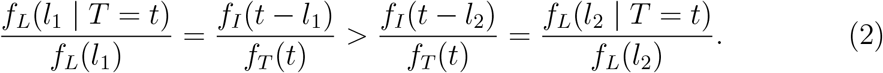

From inequality (2), we have

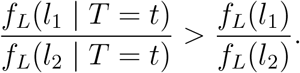

That is, conditioning on *T* = *t* shifts probability mass from larger values of *L* to smaller values. So, if we look only at those individuals whose positive test time is *t*, then small delays will be over-represented and long delays under-represented. This is not surprising, since an individual with positive test time *t* had a short delay if he was infected recently and a long delay if he was infected long ago, and there are more individuals infected recently than individuals infected long ago.

Conversely, if the incidence of infection is decreasing, *f*_*I*_(*t* − *l*) will be a increasing function of *l*, and so

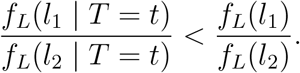

That is, conditioning on *T* = *t* shifts probability mass from smaller values of *L* to larger values: long delays are over-represented and short delays under-represented. In this situation, there are fewer individuals infected recently than individuals infected long ago.

### Example 1

Suppose half of infected individuals test positive on the day after they are infected and the other half test positive two days after they are infected. That is, *P*(*L* = 1) = *P*(*L* = 2) = 0.5. Further, suppose that 100 individuals are infected on day *t* − 2 and 150 individuals are infected on day *t* − 1 (so incidence is increasing). Then 125 individuals will test positive on day *t* and, of these, 50 were infected on day *t* − 2 and 75 were infected on day *t* − 1. So, the proportion of these 125 individuals whose delay was one day is 75/125 = 0.6 > 0.5. Conversely, suppose that 150 individuals are infected on day *t* − 2 and 100 individuals are infected on day *t* − 1 (incidence is decreasing). Then 125 individuals will again test positive on day *t*, but the proportion of these whose delay is one day is only 50/125 = 0.4 < 0.5.

### Example 2

Verity et al.[25] (see also [20]) showed that if infections are generated by a Poisson process with rate at time *t* proportional to exp(*λt*) for some *λ*, and the delay *L* has a gamma distribution with shape *α* and rate *β*, then the conditional distribution of *L* given *T* = *t* is gamma with shape *α* and rate *β* + *λ*. If *λ* > 0, then the incidence is rising (exponentially) and the conditional mean delay, *α*/(*β* + *λ*), is less than the unconditional mean *α*/*β*. If instead *λ* < 0, then the incidence is falling and the conditional mean delay is greater than the unconditional mean. If *λ* = 0, the two gamma distributions are the same.

### Example 3

Figure 1 shows how the hospitalisation risk conditional on positive test time varies according to positive test time in a scenario where the incidence of infection first rises then falls, then rises and falls again. Here, the hospitalisation risk conditional on infection time is 5% irrespective of the infection time, and the mean time from infection to positive test is shorter in individuals who are ultimately hospitalised than in those who will not.

**Figure 1:**
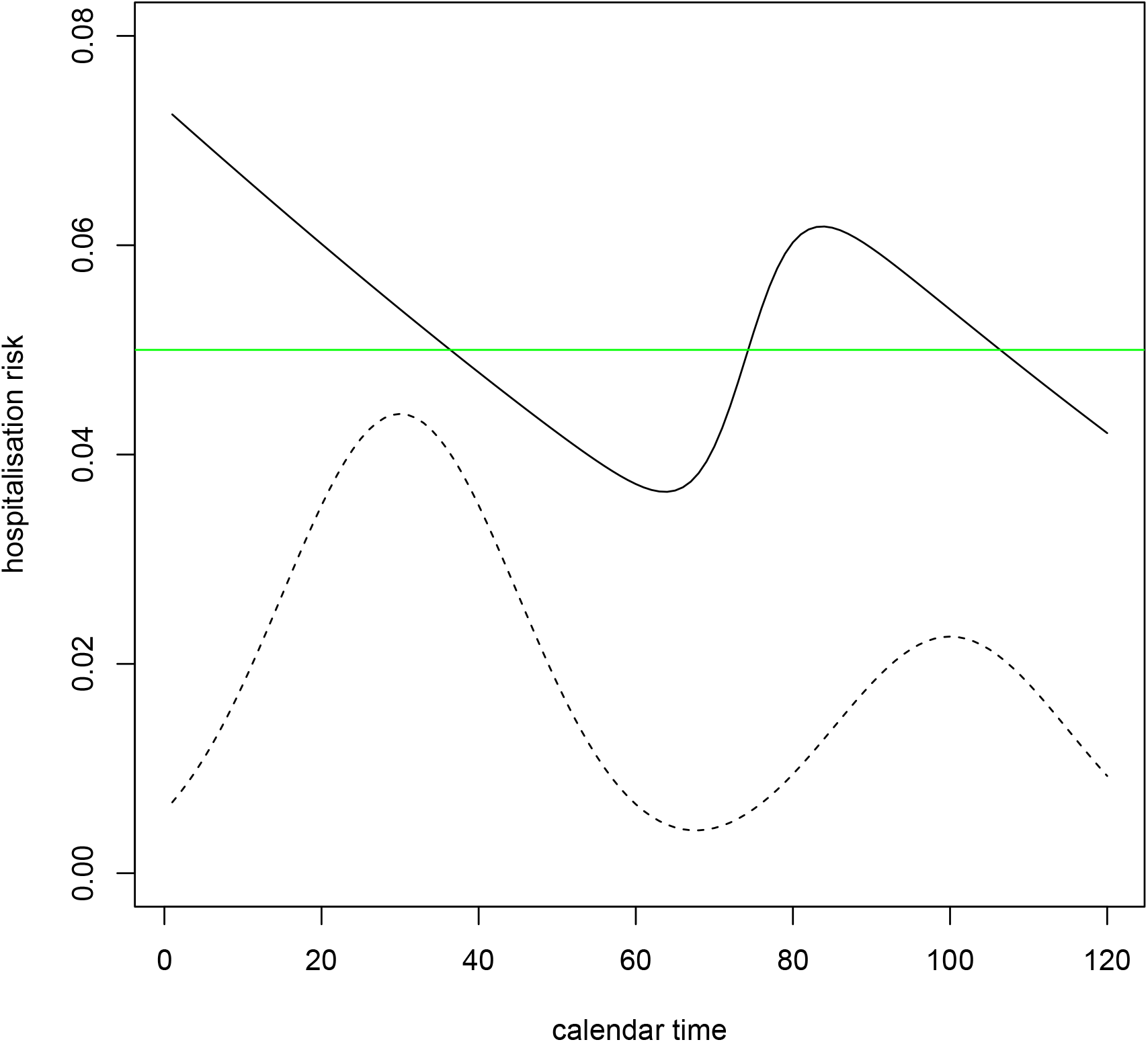
Hospitalisation risk conditional on positive test time (solid black line) when risk conditional on infection time is 0.05 (green line). Incidence of infection is shown (dotted line). Time from infection to positive test is assumed to have a gamma distribution with mean 4 and variance 8 for the ultimately hospitalised individuals and a gamma distribution with mean 7 and variance 14 for the ultimately non-hospitalised individuals.

## 4 Hospitalisation risk conditional on test time

As we have seen, conditioning on the positive test time changes the distribution of the delay in circumstances where the delay is independent of the time of infection. If hospitalisation is more common in individuals with shorter delays than in those with longer delays, i.e. *P*(*H* = 1 | *I* = *t, L* = *l*) is a decreasing function of *l*, then conditioning on the positive test time might be expected also to change the probability of hospitalisation. The effect of conditioning on *T* = *t* will depend on whether the incidence of infection is rising or falling. If it is rising, we might expect the proportion of hospitalisations to be increased, because short delays are over-represented. Conversely, if the incidence is falling, we might expect the proportion of hospitalisations to be decreased. We now confirm mathematically that this is true when the risk of hospitalisation either does not depend on the time of infection or changes little over the course of all but the longest delays.

Suppose that almost all delays are at most *l*^*\**^ for some constant *l*^*\**^ (i.e. *P*(*L* > *l*^*\**^) ≈ 0). Also, assume that *P*(*H* = 1 | *I* = *t* − *l, L* = *l*) ≈ *P*(*H* = 1 | *I* = *t, L* = *l*) for all 0 *< l* ≤ *l*^*\**^. The risk of hospitalisation conditional on time of infection can then be written as

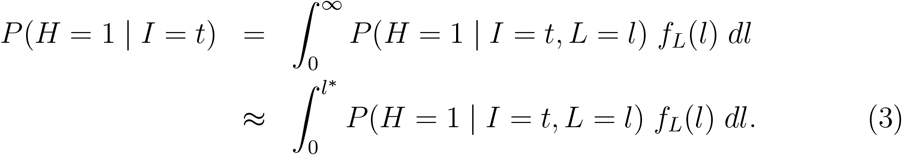

Likewise, the risk of hospitalisation conditional on positive test time is

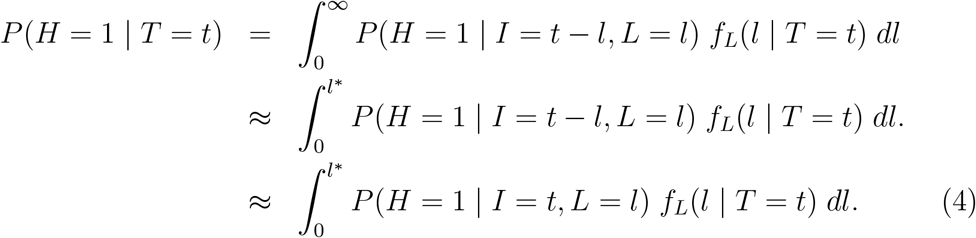

Expressions (3) and (4) are both weighted averages of *P*(*H* = 1 | *I* = *t, L* = *l*). In expression (3) the weighting function is *f*_*L*_(*l*); in (4) it is *f*_*L*_(*l* | *T* = *t*). If the incidence of infection is increasing over the period [*t* − *l*^*\**^, *t*], then, as explained in Section 3, conditioning on *T* = *t* shifts probability mass from larger values of *L* to smaller values. Hence, the weighted average in expression (4) gives more weight to small values of *l* (and less weight to large values of *l*) than does the weighted average in expression (3). This, combined with our assumption that *P*(*H* = 1| *I* = *t, L* = *l*) is a decreasing function of *l*, implies that (4) is greater than (3). That is, *P*(*H* = 1 | *T* = *t*) > *P*(*H* = 1 | *I* = *t*). On the other hand, if the incidence of infection is decreasing over the period [*t* − *l*^*\**^, *t*], then (as explained in Section 3) conditioning on *T* = *t* shifts probability mass from smaller values of *L* to larger values, with the result that *P*(*H* = 1 | *T* = *t*) < *P*(*H* = 1 | *I* = *t*).

### Example 1 continued

Suppose *P*(*H* = 1 | *I* = *t, L* = 1) = 0.05 and *P*(*H* = 1 | *I* = *t, L* = 2) = 0.01. Then *P*(*H* = 1 | *I* = *t*) = (0.05 + 0.01)/2 = 0.03. If the incidence of infection is increasing, then of the 125 individuals who test positive on day *t*, the expected number who are hospitalised is 50 × 0 0.01 + 75 × 0.05 = 4.25, corresponding to a proportion of 4.25/125 = 0.034 (which is > 0.03). If, on the other hand, the incidence of infection is decreasing, then of the 125 individuals who test positive on day *t*, the expected number who are hospitalised is 75 × 0.01 + 50 × 0.05 = 3.25, corresponding to a proportion of 0.026 (which is < 0.03). The ratio of these two proportions is 0.034/0.026 = 1.31, and so the hospitalisation risk conditional on time of positive test would differ by 31% between a period of epidemic growth and a period of epidemic decline.

## 5 Hospitalisation risk conditional on infection time plus random delay

Suppose now that we had a different proxy of infection time, which, unlike positive test time, were not associated with the hospitalisation outcome. If we conditioned on this proxy, we might achieve the goal of approximately adjusting for time of infection without creating a measure of hospitalisation risk that depends on the trajectory of the infection incidence. We now describe such a proxy.

Suppose, hypothetically, that each individual who becomes infected at time *I* = *i* and tests positive at time *T* = *t* is randomly assigned a time variable *T*^0^ sampled from the conditional distribution of *T* given *I* = *i* and *H* = 0, i.e. the distribution of positive test time in those who are infected at the same time and who are not ultimately hospitalised. This time *T*^0^ will be our proxy of infection time. By construction, it is not associated with the hospitalisation outcome *H*.

We cannot actually carry out this assignment in practice, because we do not observe *I*. However, under the following working assumption, we shall still be able to estimate *P*(*H* = 1 | *T*^0^ = *t*).

### Assumption 1

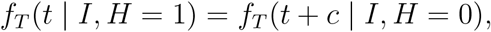

*where c is some known constant*.

Assumption 1 means that the distribution of time from infection to positive test (conditional on time of infection) in ultimately hospitalised individuals equals the corresponding distribution in ultimately non-hospitalised individuals shifted by *c* days. So, the mean time from infection to test in the ultimately hospitalised is *c* days less than the mean in the ultimately non-hospitalised. We might expect that *c* > 0.

If Assumption 1 holds, then (see Appendix A for proof)

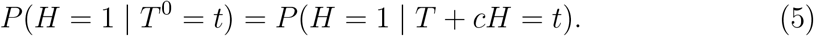

It follows from equation (5) that *P*(*H* = 1 | *T*^0^ = *t*) can be consistently estimated simply by creating the new variable *T*^*\**^ = *T* + *cH* (which equals *T* for ultimately non-hospitalised cases and *T* + *c* for ultimately hospitalised cases) and calculating the proportion who have *H* = 1 among those sampled individuals with *T*^*\**^ = *t* (or, if time is continuous, *T*^*\**^ ≈ *t*).

The hospitalisation risk conditional on *T*^0^, i.e. *P*(*H* = 1 | *T*^0^ = *t*), has the desirable property that *P*(*H* = 1 | *T*^0^ = *t*) = *P*(*H* = 1 | *I* = *t*) if *P*(*H* = 1 | *I* = *t*) does not depend on *t*. More generally, if *P*(*H* = 1 | *I* = *t*) ≈ *P*(*H* = 1 | *I* = *t* + *l*) for all 0 *< l* ≤ *l*^*\**^, then *P*(*H* = 1 | *T*^0^ = *t*) ≈ *P*(*H* = 1 | *I* = *t*). That is, when the risk conditional on *I* is constant or changes only slowly over time, conditioning on *T*^0^ yields almost the same risk as conditioning on *I*.

In Appendix B, we allow for covariates *X* that are fixed at the time of infection and for the possibility that our sample consists of individuals who are sampled conditionally on their positive test time lying within some calendar time interval, i.e. conditional on *T* ∈ [*τ*_1_, *τ*_2_], for some 0 ≤ *τ*_1_ ≤ *τ*_2_.

In practice, it is unlikely that we shall know the true value of *c*. However, one may be able to specify a range of plausible values for it and then investigate how sensitive the estimate of *P*(*H* = 1 | *T*^0^) is to this value.

Assumption 1 states that the distribution of delay in hospitalised individuals equals the distribution of delay in non-hospitalised individuals shifted by some number (*c*) of days. This assumption may well be false. In particular, it implies that the minimum delay in non-hospitalised cases cannot be less than *c*. So, in Section 6 we shall investigate the extent to which *P*(*H* = 1 | *T*^*\**^) differs from *P*(*H* = 1 | *T*^0^) when one delay distribution is not a shifted version of the other but we set *c* to be equal to *E*(*L* | *I, H* = 1) − *E*(*L* | *I, H* = 0), i.e. the difference between the mean of the two delay distributions.

## 6 Investigation of proposed method

Suppose the incidence of infection at time *t* is proportional to exp(*λt*). If *λ* > 0, then *d* = log(2)*/λ* is the doubling time; if *λ* < 0, then −*d* is the halving time. Suppose that *P*(*H* = 1 | *I* = *t*), the risk of hospitalisation for an individual who is infected at time *t* is *r* = 0.05 and does not depend on the time of infection. Suppose the delay in the non-hospitalised cases has a gamma distribution with mean 7 and variance 14 (i.e. shape *α* = 3.5 and rate *β* = 0.5). We consider two scenarios for the distribution of delay in the hospitalised cases. In Scenario 1, it is a gamma distribution with mean 4 and variance 8 (shape *α*_1_ = 2 and rate *β* = 0.5). In Scenario 2, it is an equal mixture of a gamma distribution with mean 7 and variance 14 and a gamma distribution with mean 1 and variance 2 (shape *α*_2_ = 0.5 and rate *β* = 0.5). So, in both scenarios the difference between the mean delay in the hospitalised and non-hospitalised is three days. Figure 2 shows, for each scenario, the distributions of delay in the non-hospitalised (black) and hospitalised (red) cases. The dotted line shows the distribution in the non-hospitalised cases shifted by three days.

**Figure 2:**
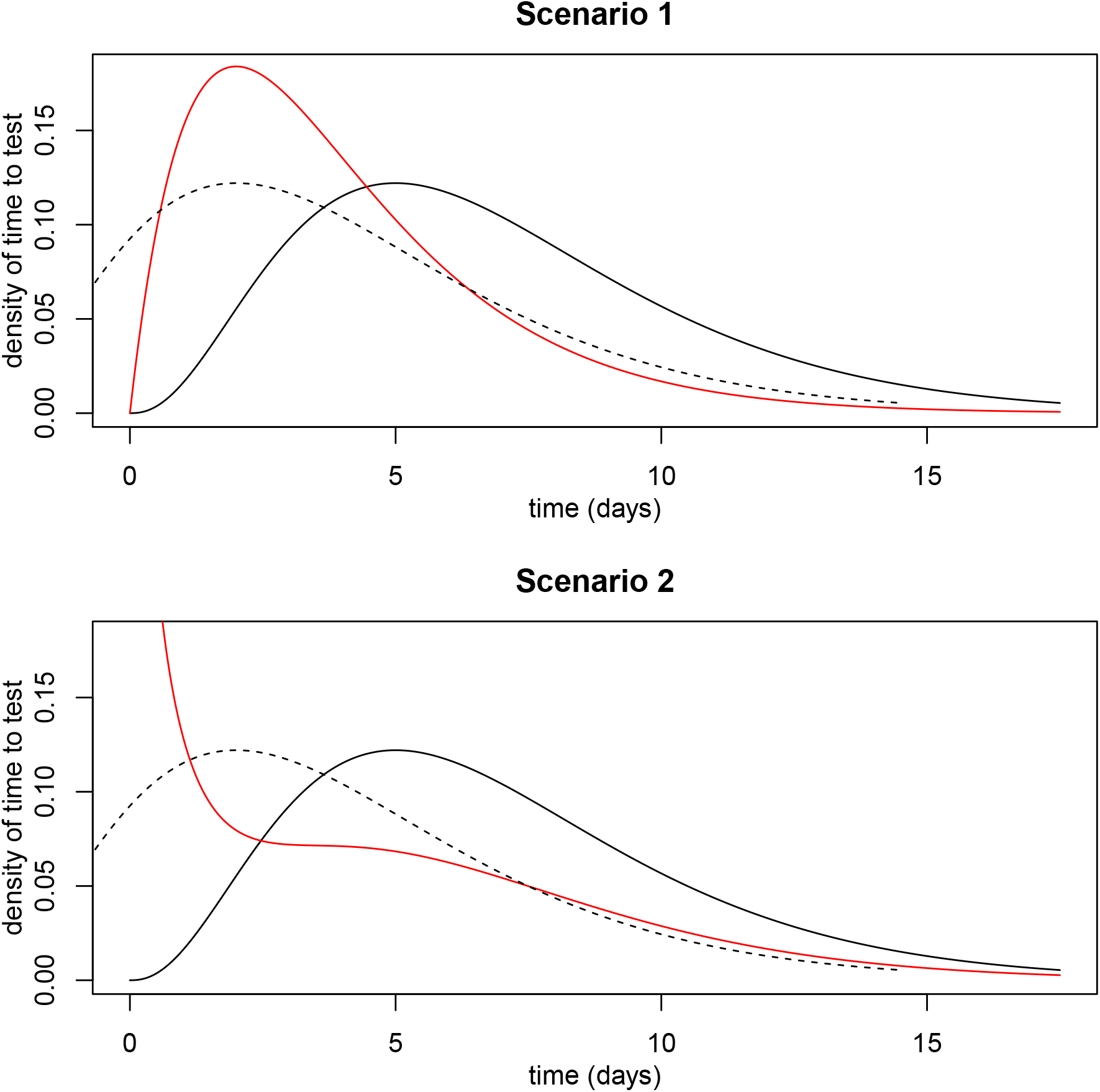
Distributions of time from infection to positive test. Solid black line is distribution for ultimately non-hospitalised individuals. Dotted line is same distribution shifted by three days. Red line is distribution for ultimately hospitalised individuals.

The number of non-hospitalised individuals who test positive at time *t* is proportional to

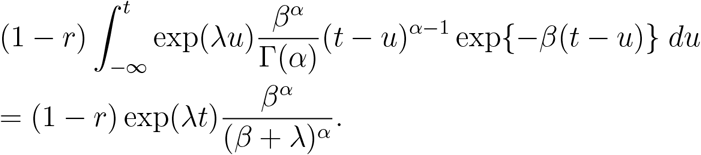

Similarly, the number of hospitalised individuals who test positive at time *t* is proportional to

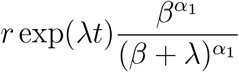

in Scenario 1 and is

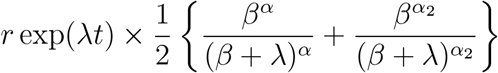

in Scenario 2.

So, the risk when we condition on time of positive test *T* is

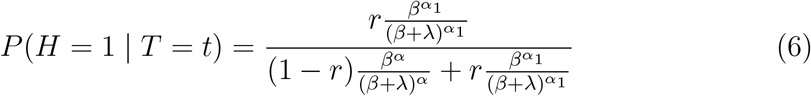

in Scenario 1 and is

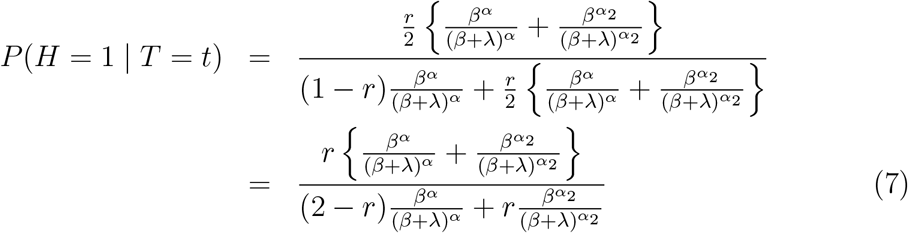

in Scenario 2.

If we condition on *T*^*\**^ = *T* + 3*H*, the risk is

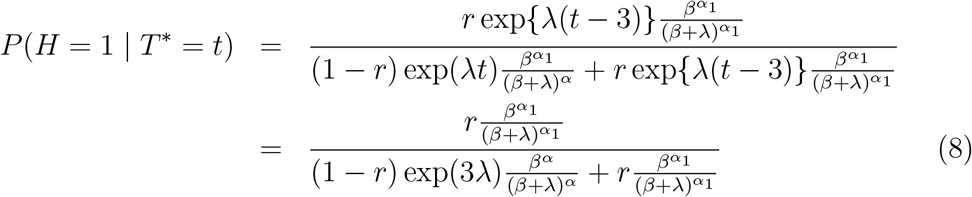

in Scenario 1, and is

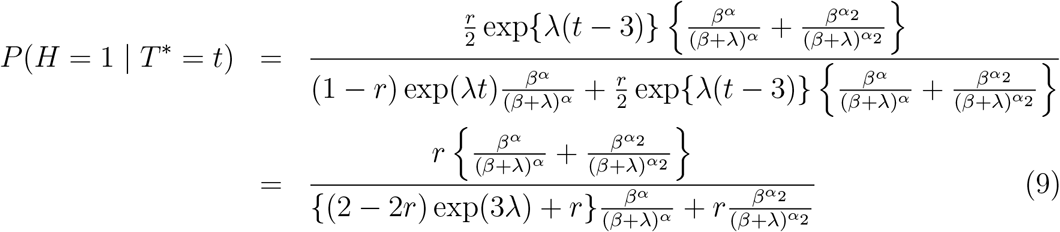

in Scenario 2.

Table 1 shows the results of applying equations (6)–(9) in Scenarios 1 and 2, when *d* = 4 and when *d* = 10. It also shows the results when *d* = − 4 or *d* = − 10, meaning that the incidence is falling with a halving time is 4 or 10 days. We see that the risks conditional on *T* are indeed different from the risk conditional on time of infection *I*, i.e. *r* = 0.05. The proposed method produces conditional risks *P*(*H* = 1 | *T*^*\**^) that are close to *r*.

**Table 1:** Risks when adjusted for *T* and *T*^*\**^. If *d* = 4 or 10, the doubling time is 4 or 10 days. If *d* = − 4 or − 10, the halving time is 4 or 10 days.

| Scenario | $d$ | $T$ | $T^*$ |
| --- | --- | --- | --- |
| 1 | 4 | 0.0760 | 0.0466 |
| 1 | 10 | 0.0601 | 0.0494 |
| 1 | -4 | 0.0270 | 0.0447 |
| 1 | -10 | 0.0404 | 0.0493 |
| 2 | 4 | 0.0830 | 0.0511 |
| 2 | 10 | 0.0612 | 0.0503 |
| 2 | -4 | 0.0326 | 0.0536 |
| 2 | -10 | 0.0414 | 0.0504 |

Finally, Table 2 shows the risk ratios that Table 1 implies when comparing two variants both of which have the same risk *r* = 0.05, but one of which has a doubling time of 4 (respectively, 10) days and the other has a halving time of 4 (respectively, 10) days. The risk ratio conditional on time of infection is *r*/*r* = 1. The risk ratios conditional on *T* vary from 1.5 to 2.8. When we instead conditional on *T*^*\**^, the conditional risk ratios vary from 0.95 to 1.04, i.e. they are much closer to 1.

**Table 2:** Risk ratios when adjusted for *T* and *T*^*\**^. One variant has doubling time *d* = 4 (or *d* = 10) days and the other has halving time *d* = 4 (or *d* = 10) days.

| Scenario | $d$ | $T$ | $T^*$ |
| --- | --- | --- | --- |
| 1 | 4 | 2.810 | 1.044 |
| 1 | 10 | 1.489 | 1.003 |
| 2 | 4 | 2.551 | 0.954 |
| 2 | 10 | 1.480 | 0.997 |

## 7 Censoring of hospitalisation outcome

So far, we have assumed that the binary outcome *H* is observed for everyone who tests positive. In practice, there may be administrative censoring. This would occur if some of the sampled individuals test positive less than 14 days before time *τ*_2_, had not yet been hospitalised by time *τ*_2_, and no data were available on hospitalisations after time *τ*_2_. In this situation, it would be natural to use the *time* from positive test to hospitalisation as the outcome (rather than the binary indicator of hospitalisation within 14 days), right-censoring this time at 14 days. Cox regression could then be used to analyse the data. When the binary outcome is rare and is fully observed, the hazard ratio (HR) estimate from Cox regression is approximately equal to the odds ratio estimate from logistic regression [1].

The proposed method now requires slight modification, because *T*^*\**^ is calculated from *T* and *H*, the last of which is unobserved for the censored individuals. To avoid compromising the simplicity of the proposed method, we suggest assuming that individuals whose hospitalisation status is unknown due to this administrative censoring have *H* = 0 for the purpose of calculating *T*^*\**^ (and so setting *T*^*\**^ = *T*). Provided that hospitalisation within 14 days is uncommon, this assumption will be true for the great majority of censored individuals.

## 8 Discussion

In this article we have highlighted the difference between the risk of a binary post-infection outcome (which, in this paper, is hospitalisation) conditional on the time of infection *I* and the corresponding risk conditional on the time of positive test *T*, and noted that the latter is a function of the trajectory of incidence of infection over calendar time. One way to interpret this difference is as a bias: if the goal is to estimate the risk conditional on infection time and if this risk differs from the risk conditional on the positive test time, then an (asymptotically) unbiased estimator of the latter risk will be an (asymptotically) biased estimator of the former risk. One might call this ‘epidemic phase bias’, since its direction and magnitude depend on whether the incidence of infection is falling or rising, and how quickly. This ‘bias’ may affect the results from a number of studies [2, 4, 5, 8, 9, 15, 16, 21, 23].

We have proposed a simple, easily implemented sensitivity analysis. This involves a third risk: that conditional on *T*^0^, the infection time plus a random time that is independent of the outcome. This third risk equals the risk conditional on infection time when the latter does not change over time (i.e. as a function of infection time), and is approximately equal to it when the risk conditional on infection time changes slowly over time. More generally, the two risks differ, but both have the advantage of not depending on the trajectory of incidence of infection.

As with other sensitivity analysis approaches, e.g. for addressing unmeasured confounding ([24]) and missing data ([7, 18]), ours does not yield a single estimate of the risk. It does, however, provide an indication of how sensitive the estimated risk is to the epidemic phase. If the incidence of infection is constant over calendar time, the estimated risk will not change as *c* is varied; if incidence is changing rapidly, the estimate will be very sensitive to the choice of *c*. When a risk ratio comparing two variants is of interest, sensitivity will be least when the variant-specific incidences of infection are both following the same trajectory (constant, increasing at the same exponential rate, or decreasing at the same rate), and will be greatest when one incidence is increasing rapidly and the other is decreasing rapidly.

The proposed method is likely to be most useful when a range of plausible values can be specified for *c*, the difference between the mean time from infection to positive test in the cases who experience the outcome and the corresponding mean time in those who do not experience the event. As a next step, we plan to apply our method to data on hospital admissions in cases infected with the Alpha and Delta variants in England, to investigate how the risk ratio changes as *c* changes.

We have allowed *c* to depend on observed covariates *X*, but have assumed it does not depend on the infection time *I* (over the study period). This assumption allows the distributions of time from infection to positive test (the ‘delay’) in the hospitalised and non-hospitalised cases to change over time, but requires that the difference between the two means remain the same. In practice, however, if both distributions are, for example, getting shorter over calendar time, then it is likely that the difference between their means will also get smaller. If it were necessary to allow *c* to depend on the unknown infection time *I* over the study period, then a crude but practical way of doing this would be to specify *c* as a function of *T* and calculate *T*^*\**^ as *T*^*\**^ = *T* + *c*(*T*).

We have focused on an observed binary outcome *H*, but also briefly addressed right-censoring of this outcome. We proposed that *T*^*\**^ be calculated as though the censored individuals did not experience the outcome (i.e. *H* = 0). This is a reasonable approximation when the outcome is rare and the proportion of censored individuals is small. For more common outcomes or when the extent of censoring is larger, it would be preferable to use a more sophisticated approach. More research is needed on this, but one possibility may be the following. Fit the Cox model to the original data. Estimate the baseline hazard. Use this estimated baseline hazard and the estimated hazard ratios from the Cox model to calculate the probability *p*_*i*_ that a censored individual *i* has *H* = 1. Then create two copies of each censored individual *i*: one with *H* = 1, *T*^*\**^ = *T* + *c* and weight *p*_*i*_; and one with *H* = 0, *T*^*\**^ = *T* and weight 1 −*p*_*i*_. An obvious drawback of this method is that *p*_*i*_ would be calculated from a model that implicitly assumed *c* = 0. A more refined version might begin by calculating *T*^*\**^ as though the censored individuals all had *H* = 0, then using the resulting fitted Cox model and estimated baseline hazard to calculate *p*_*i*_.

We have assumed that all infections result in a positive test. This is obviously not true in reality. However, this issue affects all studies of risks of post-infection outcomes in samples of individuals who have tested positive, and is not specific to this article. There is not a problem if those individuals who test positive are representative of all infected individuals. Otherwise, the estimated risks must be interpreted as risks conditional on eventually testing positive.

Finally, if additional information is available on the incidence of infection with each variant over time, it may be possible to estimate the hospitalisation risk without using data on positive test times. This could be done using deconvolution techniques, such as those developed in the 1980’s and 1990’s for back-calculation in the context of the HIV/AIDS epidemic. There the purpose was to estimate the distribution of HIV infection times from the observed distribution of AIDS onset times and an assumed-known distribution of time from infection to AIDS onset. For example, Rosenberg and Gail (1991) described how to do this using software for Poisson regression with identity link function [17]. In the context of the present article, the purpose would be to estimate, for each variant, the distribution of time from infection to hospitalisation from the observed distribution of hospitalisation times and an assumed-known distribution of infection times. It may be possible to do this by applying, for example, an adaptation of the Poisson regression method of Rosenberg and Gail with an additional offset term for the total number of infections observed so far.

## Data Availability

Not applicable; no data presented.

## Appendix A: Proof of equation (5)

The probability distribution function of the observed data (*X, T, H*) at (*x, t, h*) is

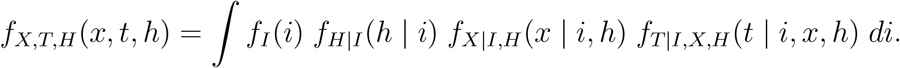

By definition, *T*^0^ is sampled from

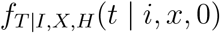

independently of *T*. So, the joint probability distribution function of (*X, T*^0^, *H*) evalulated at (*x, t, h*) is

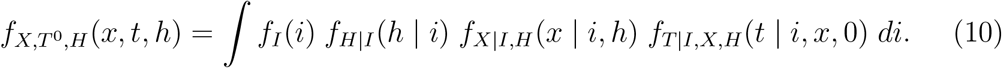

Let *T*^*\**^ = *T* + *cH*. Then the joint probability distribution function of (*X, T*^*\**^, *H*) evalulated at (*i, t, h*) is

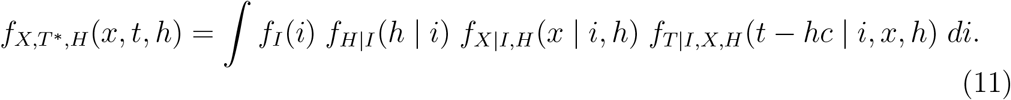

Assumption 1 can be written as

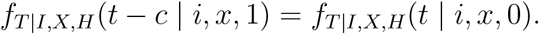

So, equation (11) becomes

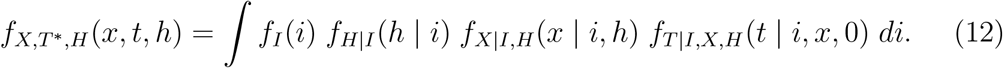

The right-hand sides of lines (12) and (10) are the same. So, (*X, T*^*\**^, *H*) and (*X, T*^0^, *H*) have the same joint distribution. Therefore, the conditional distribution of *H* given *X* and *T*^0^ is the same as the conditional distribution of *H* given *X* and *T*^*\**^.

## Appendix B: covariates and sampling within a time interval

Here we extend the results of Section 5 to allow for covariates *X* that are fixed at the time of infection and for the possibility that our sample consists of individuals sampled conditionally on *T* ∈ [*τ*_1_, *τ*_2_], for some 0 ≤ *τ*_1_ ≤ *τ*_2_.

We now replace Assumption 1 with the following more general assumption.

### Assumption 2

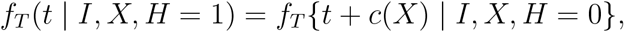

*for all t* ∈ [*τ*_1_, *τ*_2_] *and for some function c*(*x*) *of x*.

In practice, we would often assume that *c*(*x*) = *c* is a constant. Assumption 2 implies that

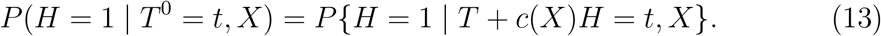

The proof that equation (13) follows from Assumption 2 is a minor generalisation of the proof given in the Appendix A.

Now we redefine *T*^*\**^ slightly as *T*^*\**^ = *T* + *c*(*X*)*H*. Because we have sampled conditionally on *T* ∈ [*τ*_1_, *τ*_2_], we have *T*^*\**^ ∈ [*τ*_1_, *τ*_2_] for all sampled individuals with *H* = 0 and *T*^*\**^ ∈ [*τ*_1_ + *c*(*x*), *τ*_2_ + *c*(*x*)] for all sampled individuals with *H* = 1 and *X* = *x*. So, assuming that *c*(*x*) ≥ 0, we can only estimate *P*(*H* = 1 | *T*^0^ = *t, X* = *x*) for *t* ∈ [*τ*_1_ + *c*(*x*), *τ*_2_]. All individuals with *H* = 0 and *T < τ*_1_ + *c*(*X*) and all individuals with *H* = 1 and *T* > *τ*_2_ − *c*(*X*) will have *T*^*\**^ values that lie outside the interval [*τ*_1_ + *c*(*X*), *τ*_2_], and hence should be dropped from the data set. One can then fit a regression model for *P*(*H* = 1 |*T*^*\**^ = *t, X* = *x*) to the remaining data. For example, if *X* = (*V, U*), where *V* is an indicator of variant and *U* is a vector of age and ethnicity, then we might fit the model

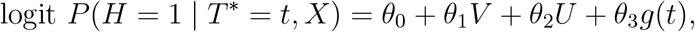

for some specified function *g*(*t*) of *t*. Now, *θ*_1_ can be interpreted as the log odds ratio of hospitalisation associated with variant adjusted for age, ethnicity and *T*^0^.

